# Reproductive health needs of Human papillomavirus (HPV) Positive women: A systematic review

**DOI:** 10.1101/2022.03.29.22273127

**Authors:** Mina Galeshi, Hoda Shirafkan, Shahla Yazdani, Zahra Motaghi

## Abstract

**Background:** Human papillomavirus is one of the most important transmitted viruses that causes of cervical cancer.

In women undergoing cervical screening, it is important to be aware of ways to reduce anxiety. The present systematic review conducted to determine the reproductive health needs of women with HPV.

**Methods:** In this systematic review, articles without time constraints searched in PubMed, Scopus, Web of Science, Google Scholar and Iranian Magiran, SID and Iranmedex.

Keywords used: HPV, Information, Want, Need, Know, etc. and their Persian equivalents in the title and abstract of the articles.

Papers after identification by two researchers Contradictions evaluated and discussed with the third author.

**Results:** At first 1056articles retrieved which after removing,13articles published between2004-2021were entered. The studies were qualitative(N=9),quantitative(N=3), and one was unclear.Most qualitative studies collected data using individual interviews(N=7), two qualitative studies, narratives of HPV patients from a website of patient experiences and questions. The quality evaluation showed that good=8, Average=2, and one was of poor. There is not enough information about Friedman AL etal and Garland SM articles to check the quality, but since these studies were valuable, they included in the study according to the opinion of the research team.

**Conclusion:** Surveys showed that the majority of women had unanswered questions about their HPV test results. The information that women thought was helpful in interpreting their test results included having a high-risk type of HPV, the risk of short-term and long-term cancer, and cancer survival statistics for the virus. Women also needed information about sexual transmission, how HPV tested positive in a long-term relationship, and the potential consequences for their partners and the risk of re-infection. Younger women had questions about whether HPV could affect fertility.

## Introduction

Human papillomavirus (HPV) is one of the most important sexually transmitted viruses (1). This virus is one of the causes of cervical cancer (2). Although this cancer has many environmental and genetic causes (such as smoking, early marriage, high parity, etc.), the most important cause is infection with HPV (3). Cervical cancer is the second most common cancer in women, with 500,000 new cases reported each year (4). Various and extensive studies have been conducted to investigate the prevalence of HPV infections in different countries as well as in Iran, which indicates its increasing prevalence in the world (5-7). The prevalence of this virus in the world is between 10.5-55.4% and in Iran, it is 49.5% (6, 7). The prevalence of genital HPV infection in the United States among women aged 18 to 59 years was 40% for all types of HPV and 20% for high-risk HPVs (8).

In general, cervical cancer is both preventable and treatable, and its prevalence is 6 times higher in developing countries than in developed countries, mainly due to differences and the lack of appropriate screening programs in these countries (9). More than 95% of cervical cancers are attributed to the papillomavirus (10). More than 100 strains of the virus have been identified, but types 16 and 18 are the most important strains of the virus in developing cervical cancer (1). Types 6 and 11 of this virus cause warts in the vulva (11), vagina, cervix, and penis (12).

This disease affects the quality of life of individuals and imposes financial costs on health systems (13). It has been found that 35-65% of people become infected with HPV during sex (14). Understanding cervical cancer, along with being involved with HPV infection, raises questions about needs and preferences. In women undergoing cervical cancer screening, it is important to be aware of ways to minimize anxiety. Previous research has been more limited to the field of HPV DNA testing (15, 16). HPV DNA testing included information on HPV disease, prevention, treatment, and risk of cancer (17).

Having an HPV test and getting the result can cause undesirable psychosocial reactions. To prevent anxiety and psychological distress, education and counseling are increasingly important for women diagnosed with HPV. Therefore, more attention should be paid to how to communicate and provide information (18-20).

Information challenges and communication gaps for women with HPV in the stages of cervical cancer screening in primary care, waiting time to refer to a specialist, first consultation, and Occurs after consultation in specialized care. Relatively little research has shown that health care providers have knowledge gaps and there is no adequate flow of HPV information between health care providers and women with HPV (21, 22).

Counseling with HPV patients poses special challenges for physicians and patients, especially in communities where HPV is rare and associated with severe stigma. It will be invaluable to provide a deep understanding of the challenges, perspectives, experiences, and needs of women living with HPV positively about healthcare providers at the primary and specialist care levels. Therefore, the present systematic review was performed to determine the needs of women with HPV.

## Material and Methods

The present study was approved by the Ethics Committee of Shahroud University of Medical Sciences: IR.SHE.REC.1400.154 and after receiving the code from the Prospero system Code: CRD42021293223 Based on the proposed systematic review and meta-analysis checklist (PRISMA) was done.

### Search strategy for identifying papers

Related articles, with electronic search in medical databases such as: PubMed, SCOPUS, Web of Science, Google Scholar search engine and Iranian Magiran, Scientific Information Database and Iranmedex and additional articles with a gray literature search using OpenGrey (www.opengrey.eu)were identified. Without language restrictions and the date of publication of the article until December 2021 was reviewed.

Keywords used included: (HPV Human Papillomavirus), Alphapapillomavirus, (Human Papillomavirus, HPV), Information, Want, Need, Know, Qualitative Research, theoretical framework, Comparative Analysis, and their Persian equivalents and the combination of keywords with or, and in the title and abstract of the articles.

### Selection process

As the reproductive health needs of women with HPV were not fully understood, a comprehensive and systematic study was conducted on the subject. Inclusion criteria include Studies related to the needs of women with HPV, Conference-related abstracts, and Exclusion criteria: Studies related to HPV in men, Complications of HPV, HPV-related risk factors, cervical cancer, Treatment of cervical cancer or colposcopy, commentaries, opinion pieces, and editorials.

### Data extraction

All the articles obtained in the introductory search at first reviewed by title, then Abstract, and finally their full text. The articles were reviewed by two people independently. The names of the authors and the journal were not hidden from reviewers. Disagreements between reviewers were decided by negotiating with a third reviewer until a final agreement was reached. The data extracted for analysis including the author’s name, year of the study, place of the study, study method, and the sample size in each group were entered into the electronic datasheet. Researchers contacted their authors for more information when they could not retrieve articles from authorized databases. All studies were reviewed for duplication. For each study, the data is listed in the table based on the study checklist.

### Quality assessment

A quantitative/qualitative adjusted checklist (CASP) was used to critically evaluate the articles. The Checklist (CASP) is a standard tool for evaluating articles, developed by the JAMA Group in 1994, and is the oldest and most widely used critical evaluation tool for a variety of articles/studies (23).

The present checklist was 10 items to help understand qualitative research. How to use this tool for evaluating qualitative studies is as follows: When evaluating, three general issues should be considered first. A: Are the study results valid enough?, B: What are the results? A: Will the results help locally? The first two questions are systematically screening questions and can be answered quickly. If the answer to both is “yes”, the remaining questions are continued each item was assigned a score of 2 (including that item in the article) or zero (not paying attention to the item in the article) and one (cannot be said) and the total score of this checklist was between 0-20.

To evaluate cross-sectional studies, this tool consisted of 18 items, and each item was given a score of one. Including that item in the article (or zero) was considered as not paying attention to the item in the article. Attitude evaluation (3 items), study design (5 items), and results (5 items) were divided and the total score of this checklist varied between 0-18.

After carefully reading the full text of each article, an article quality evaluation checklist was completed by the first researcher and items were scored. Reassessment was performed in the same way by the second researcher. If there was no agreement on scoring the items, the final score would be taken in a joint session.

In the next step, these articles were compared in terms of scores obtained in each of the four areas as well as in terms of total scores. Also, the qualitatively reviewed articles were divided into three categories of good, medium, and poor quality articles according to the scores obtained from this checklist. Total score 20 high quality; 16-19 Medium quality; ≤15 was considered low quality, also the articles reviewed cross-sectional in consultation with experts, scores of 75% of the total score and above as good quality (score≥ 13), scores between 25-25% of the total score as average quality (scores 6-12) and scores less than 25% of the total score (score 5 and below) were divided as poor quality (24, 25).

### Analysis

Quantitative and qualitative findings were analyzed separately. The results of quantitative studies were reported descriptively by first creating an initial combination of findings, existing relationships and data strength were examined.

For qualitative studies, a thematic synthesis was performed by examining the text line by line in the results section and discussing and then identifying descriptive topics.

## Results

In the search of databases, at first 1056 articles were retrieved, which after removing 963 duplicate and unrelated articles, 61 articles were evaluated based on inclusion criteria (Flowchart).

Studies were conducted in the United States (N=5), Europe (N=4), Asia (N=4), and published between 2004-2021. The studies were mostly qualitative (N=9), the study was quantitative (N=3), and one type of study was unclear. Most qualitative studies collected data using individual interviews (N=7), two qualitative studies, narratives of HPV patients from a website of patient experiences and questions. The characteristics of the participant and the study are shown in the table (Table 1).

**Table1:**
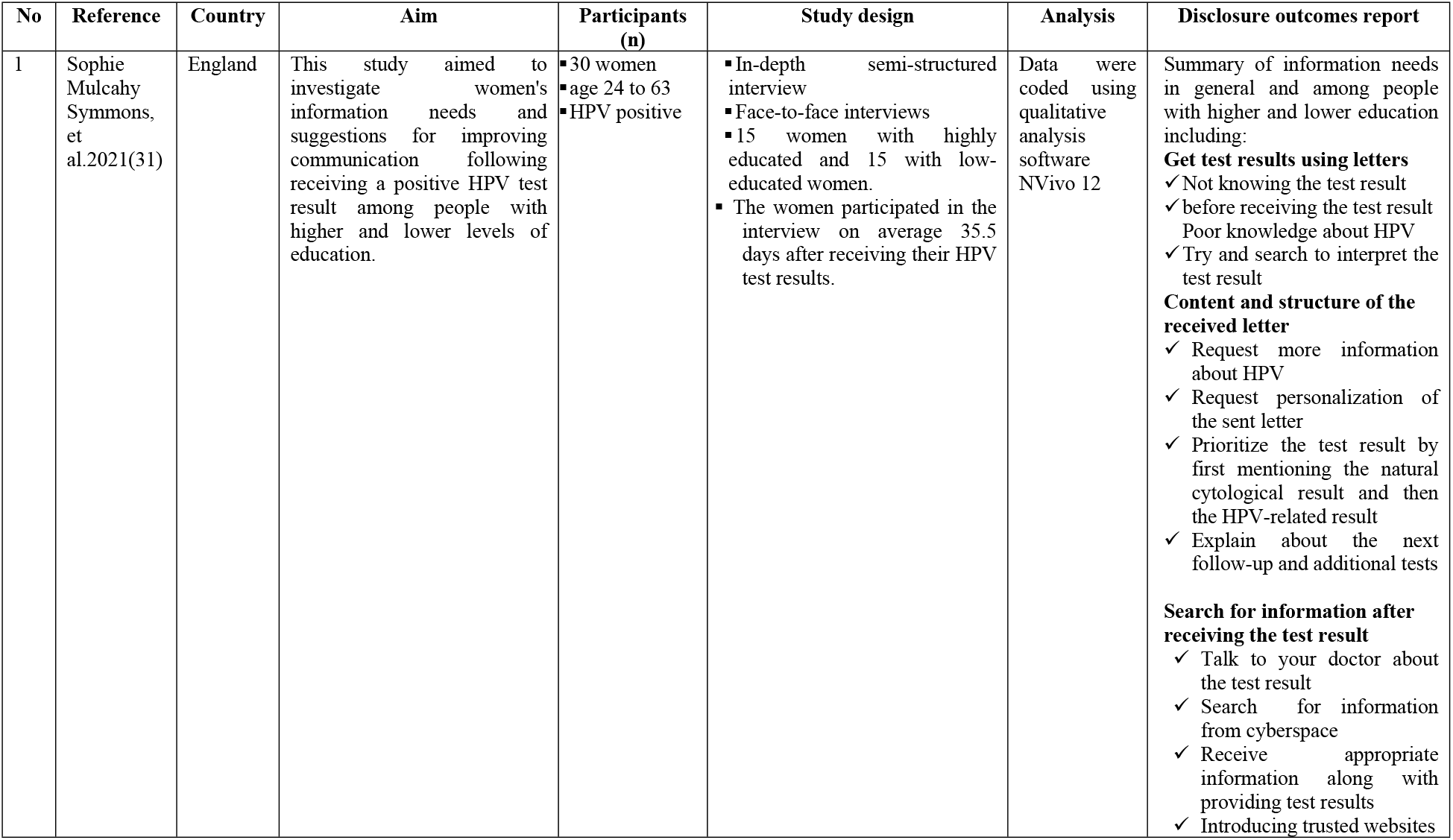
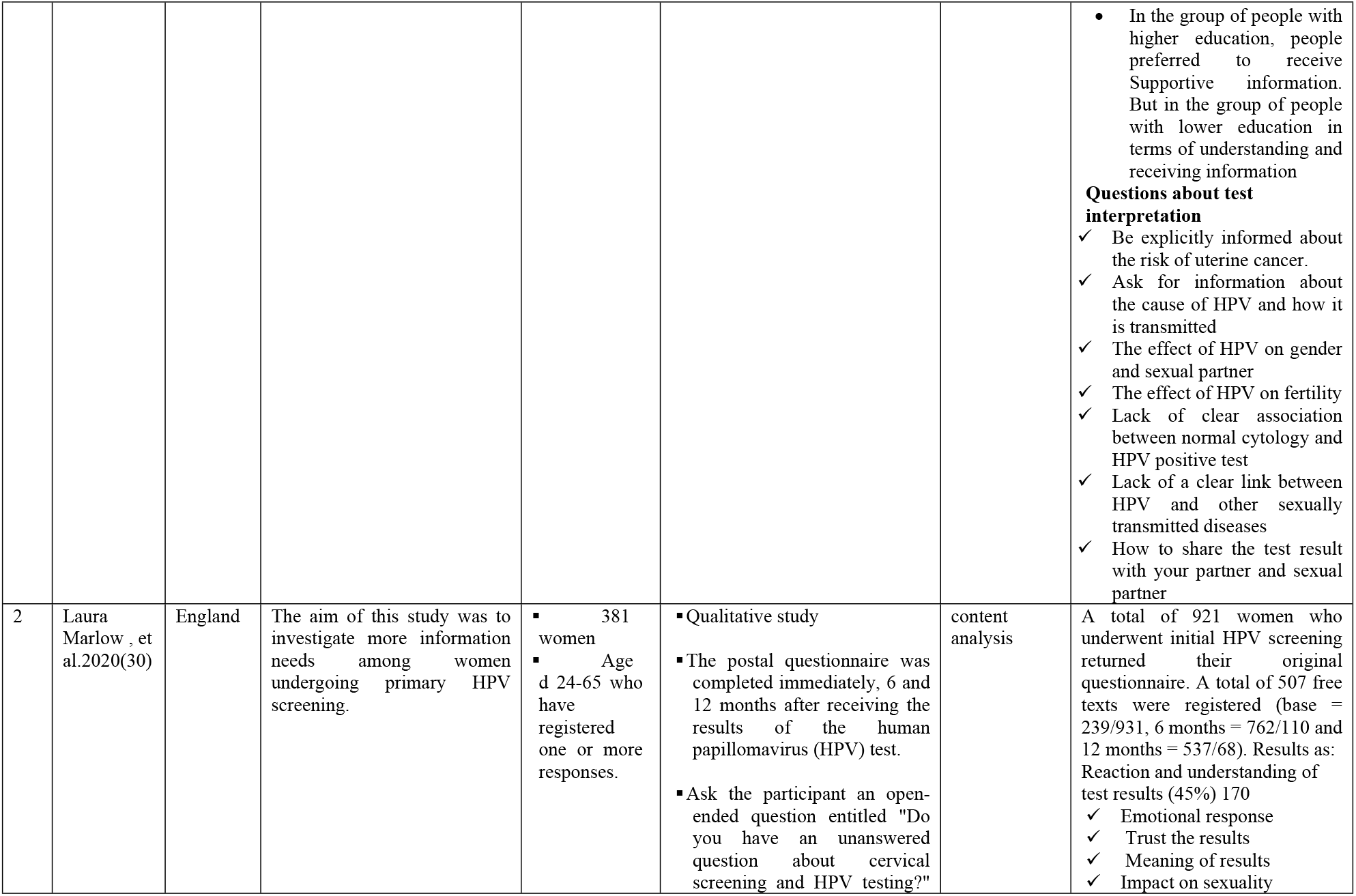
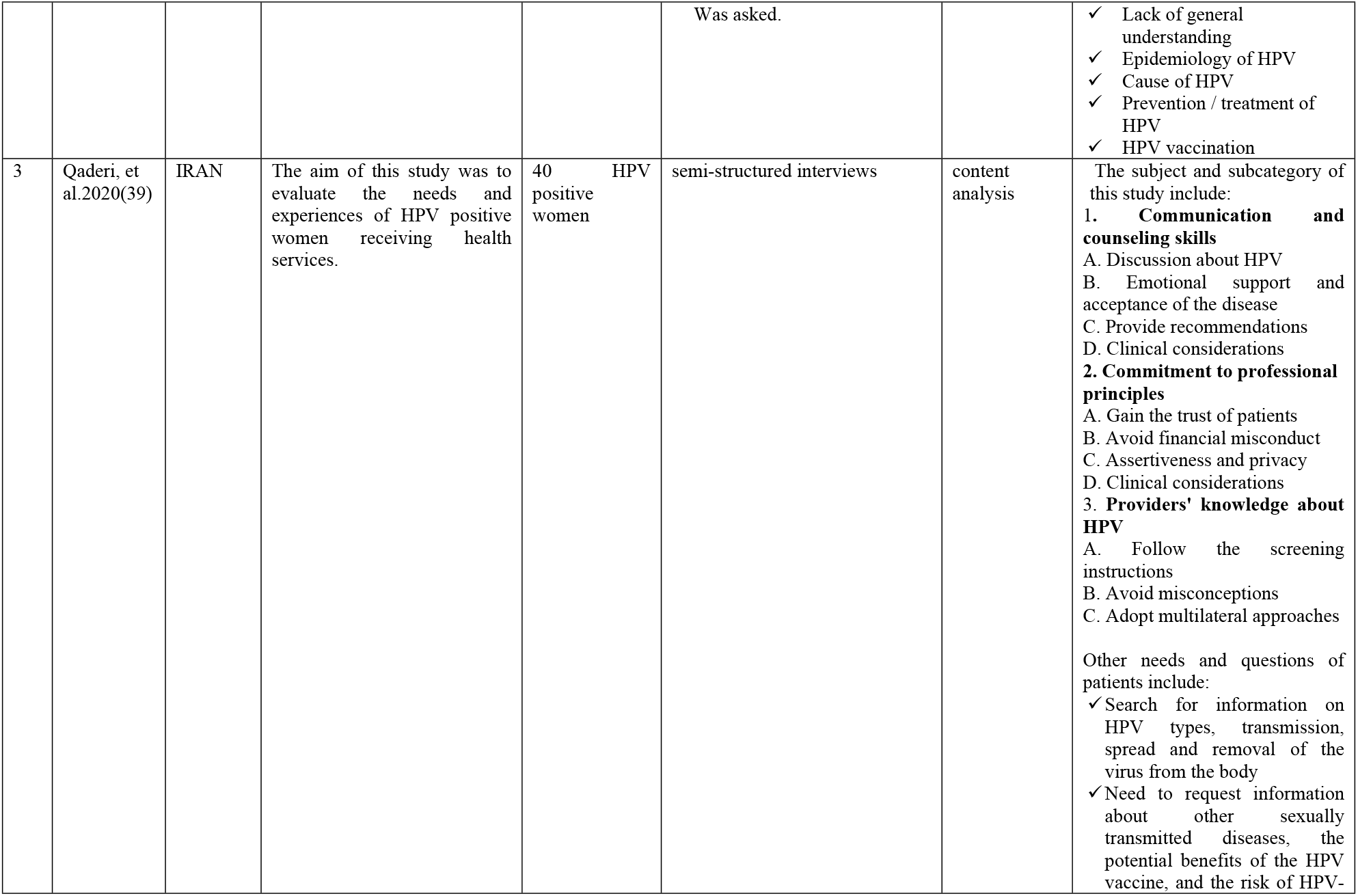
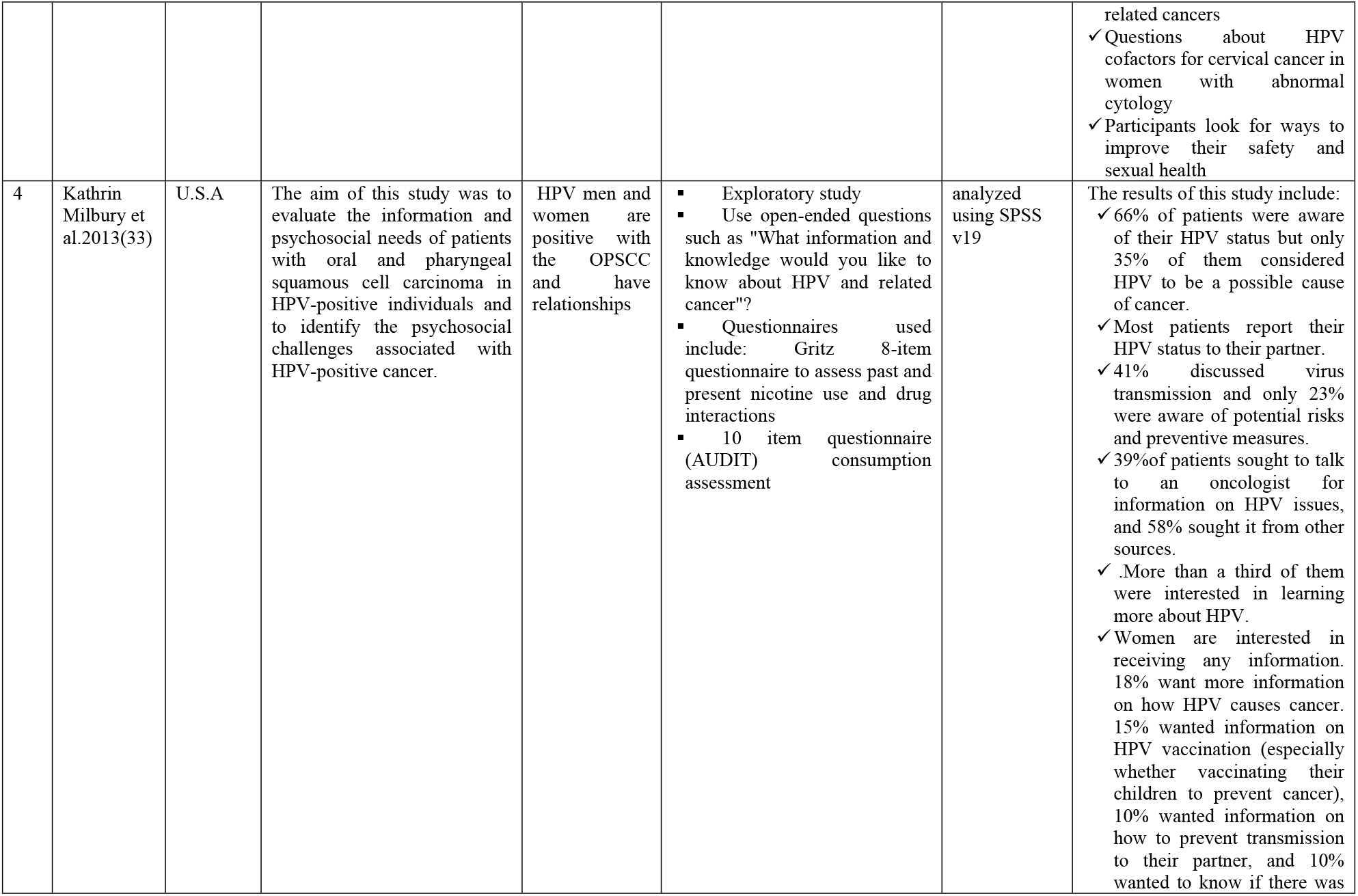
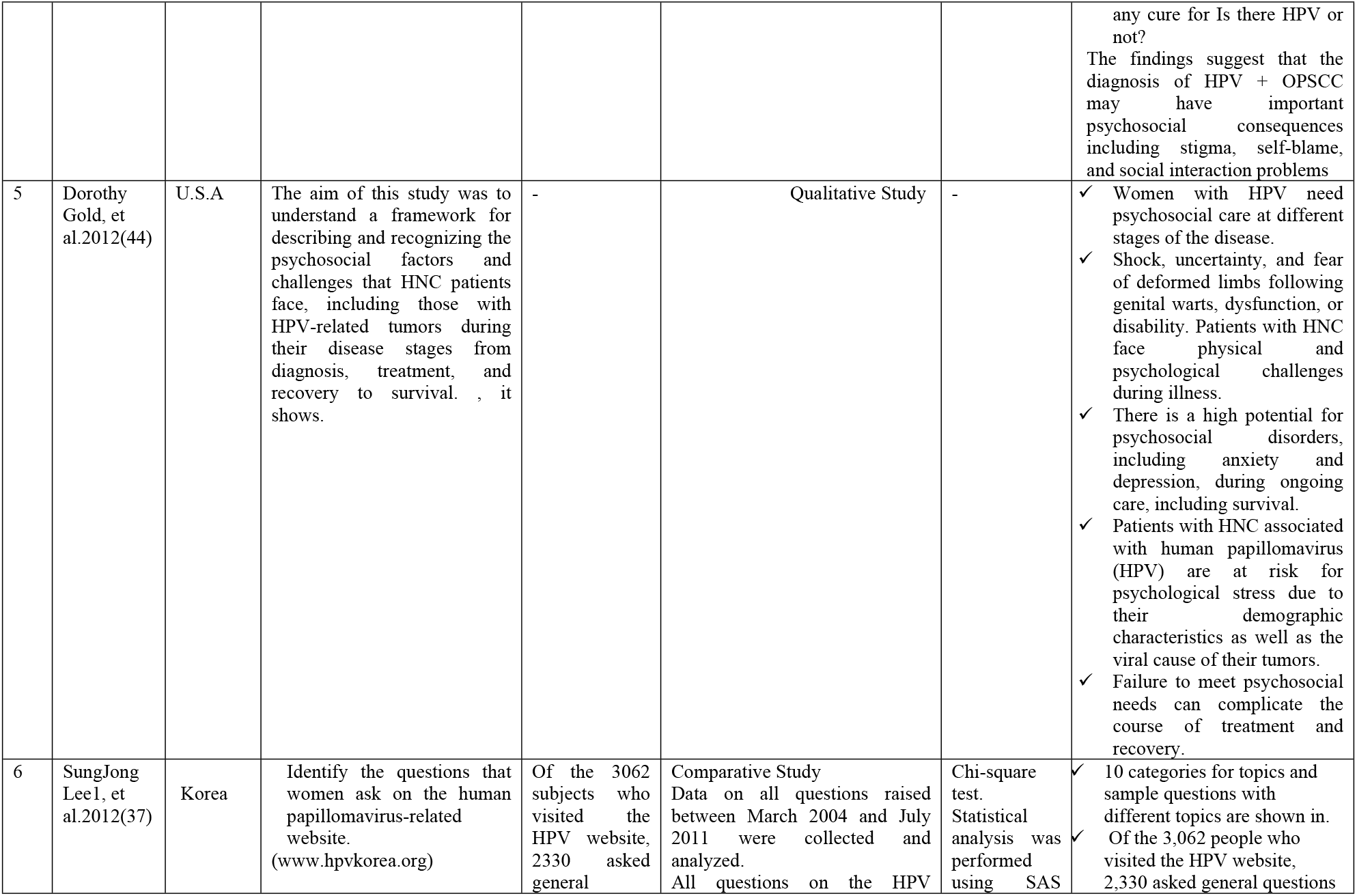
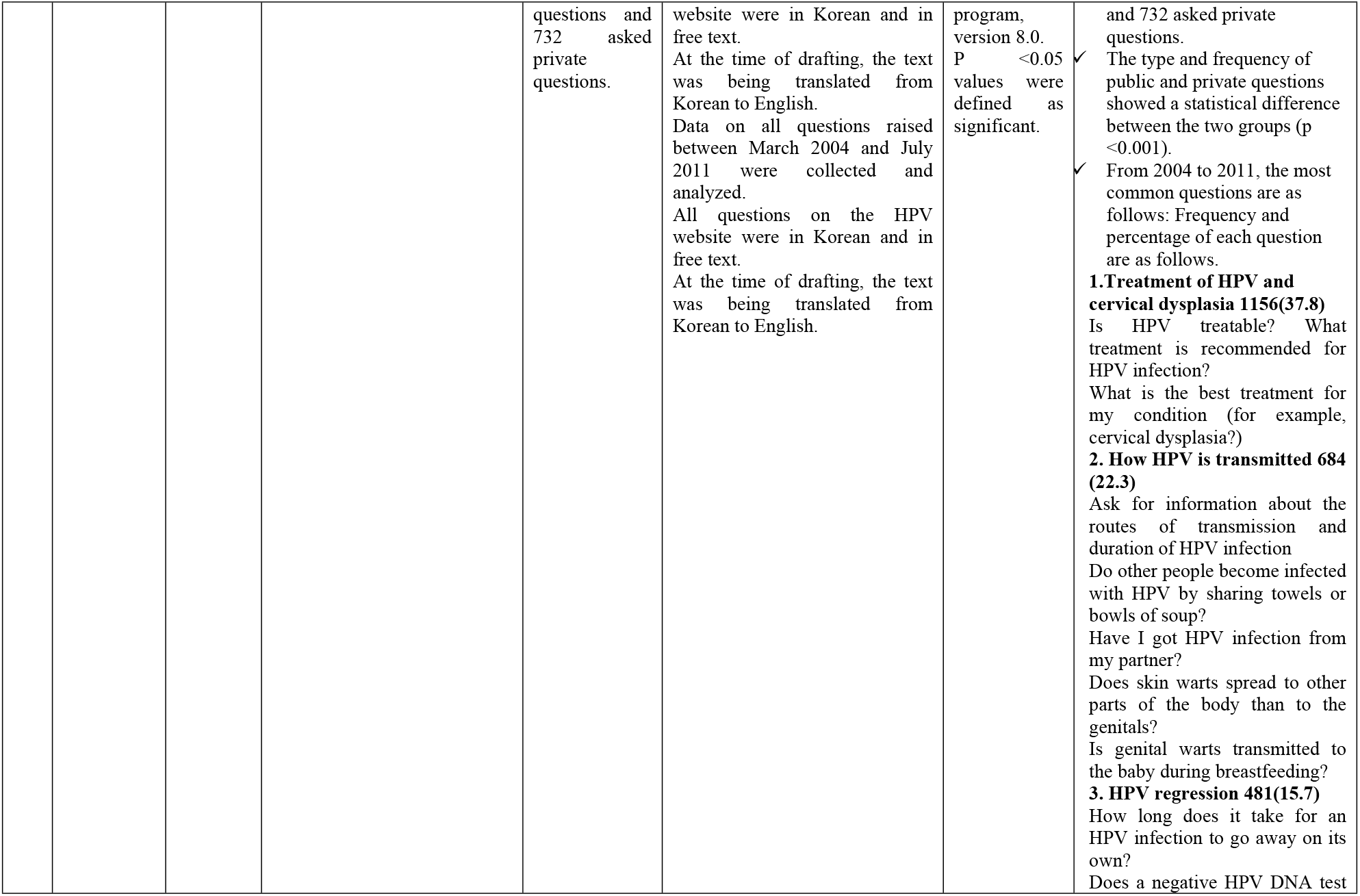
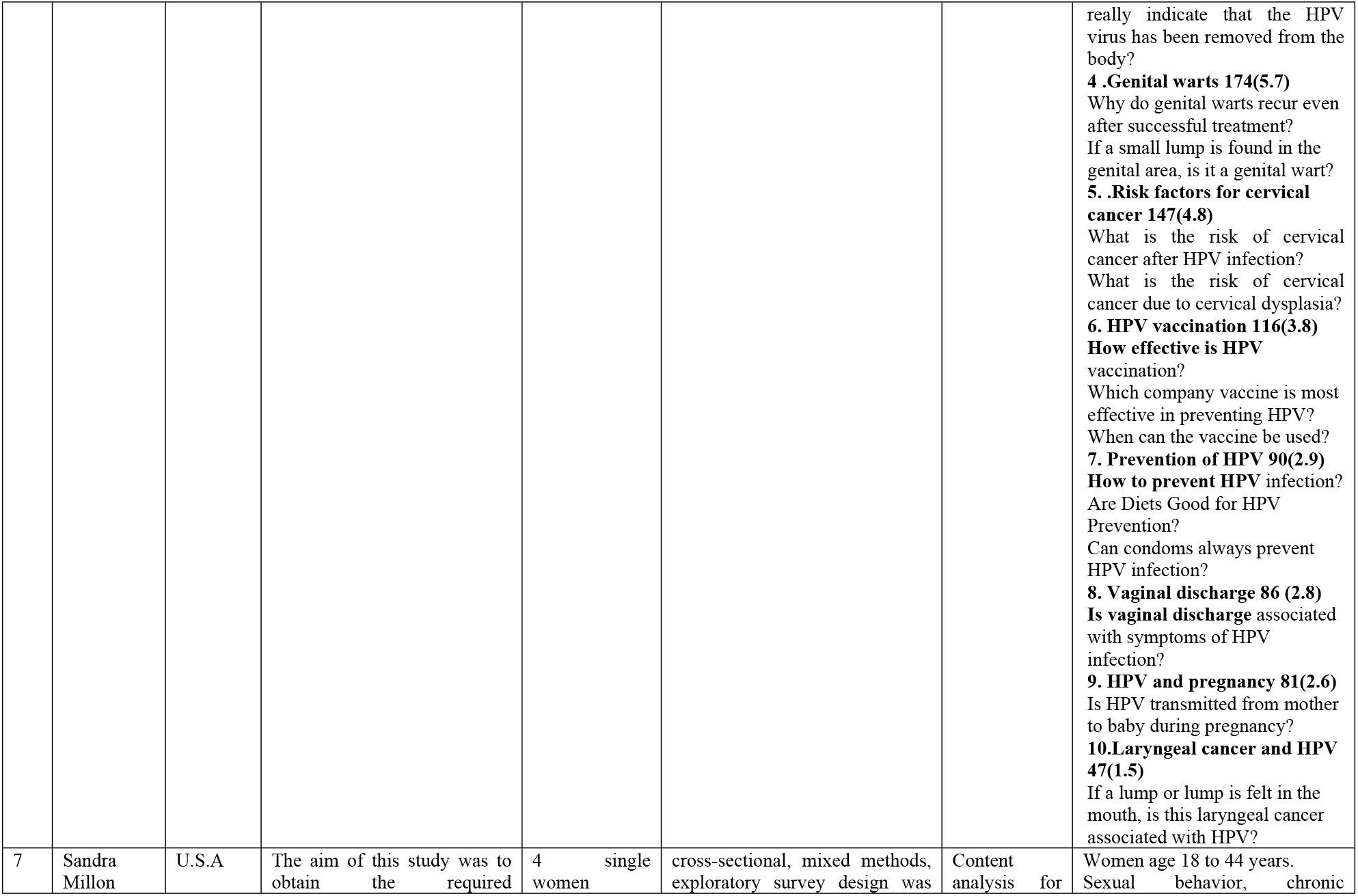
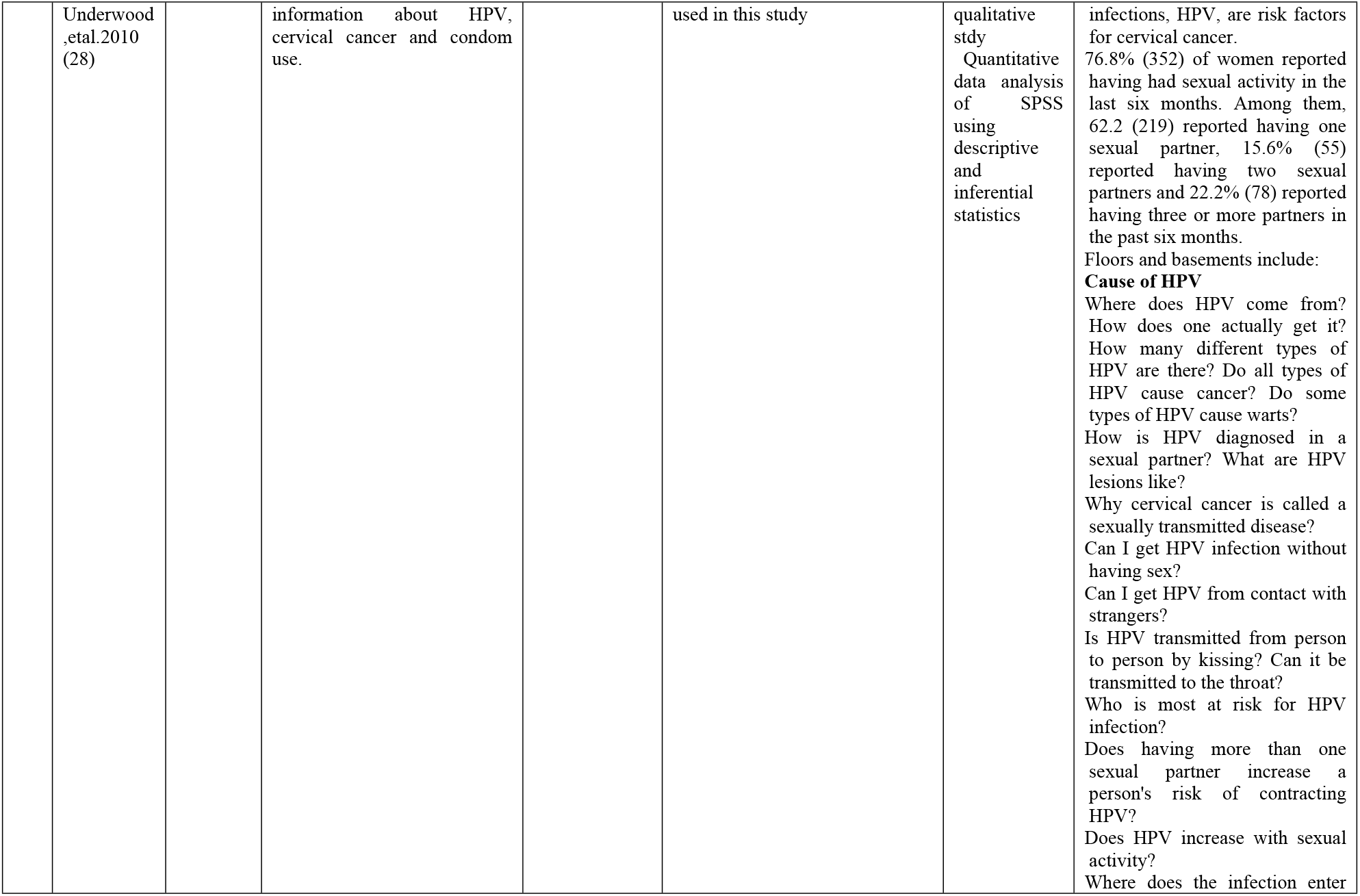
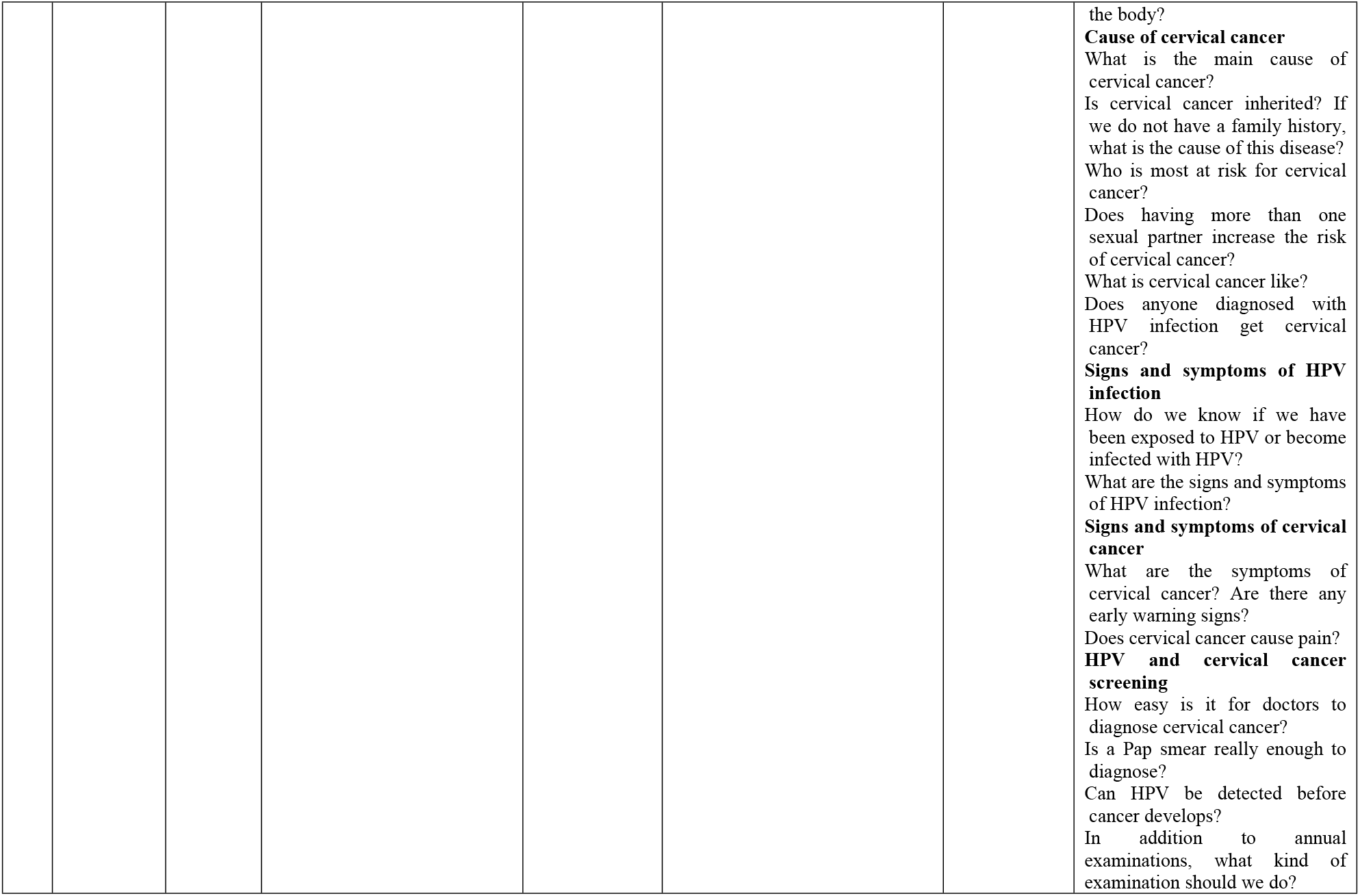
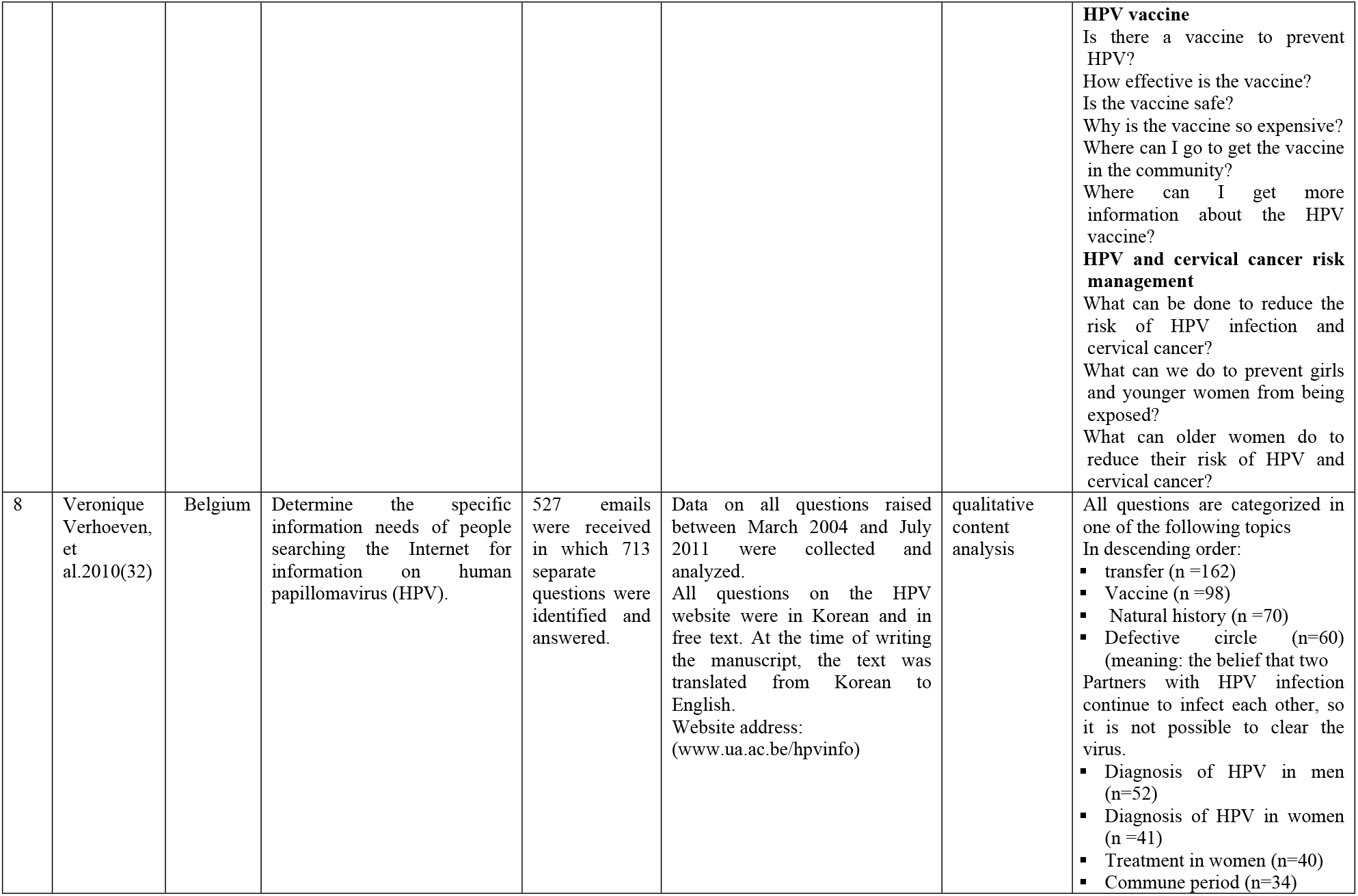
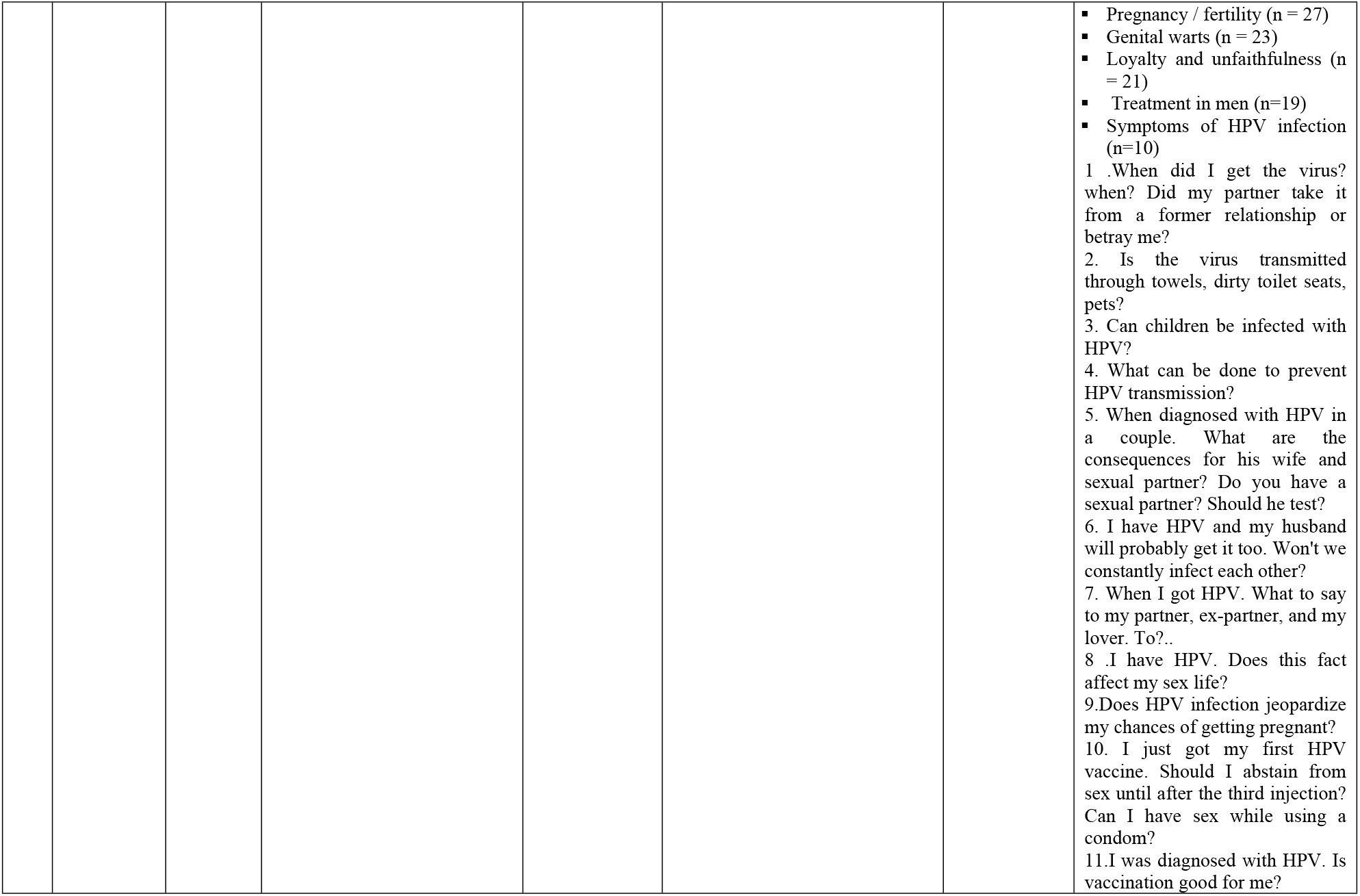
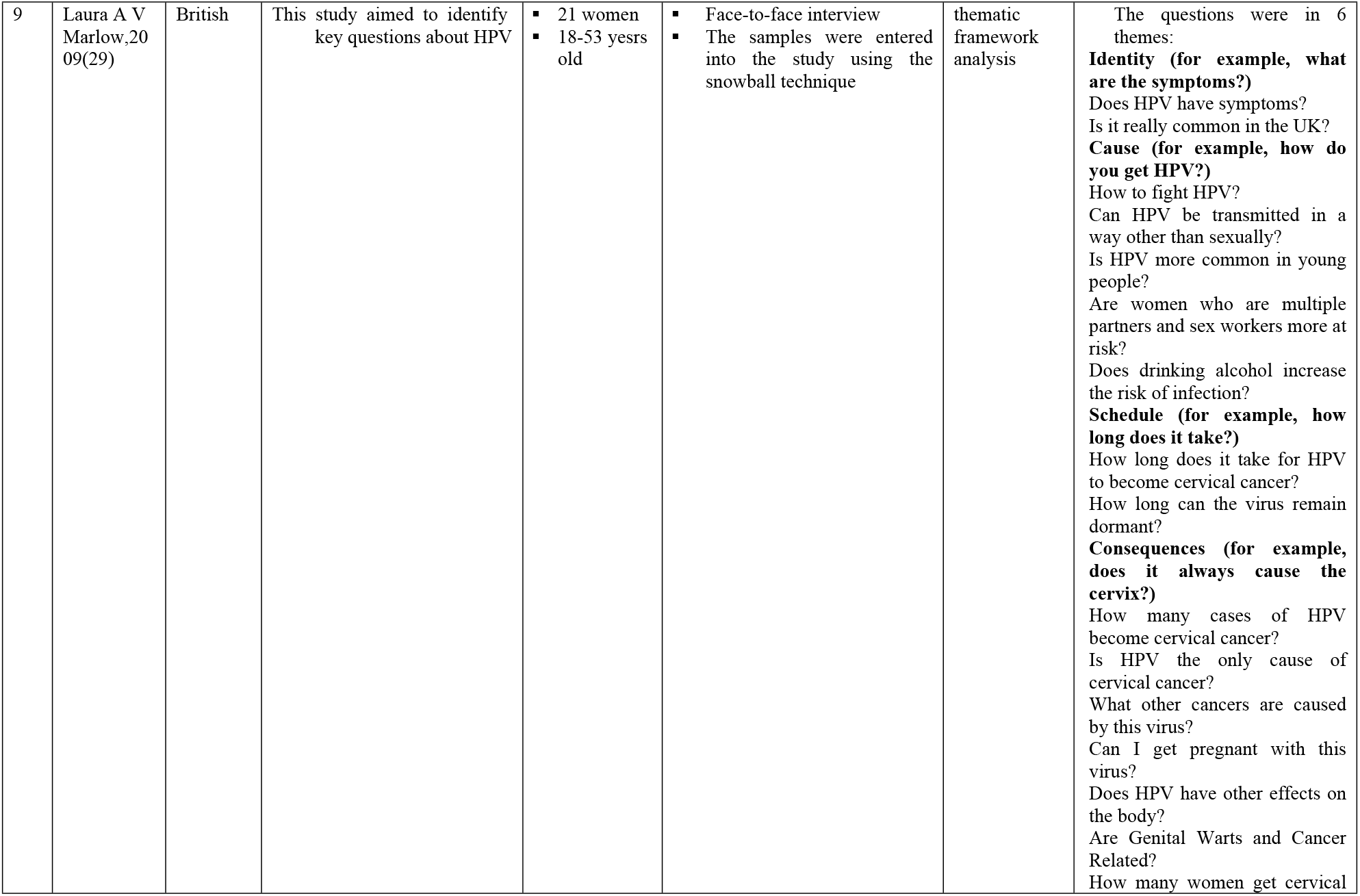
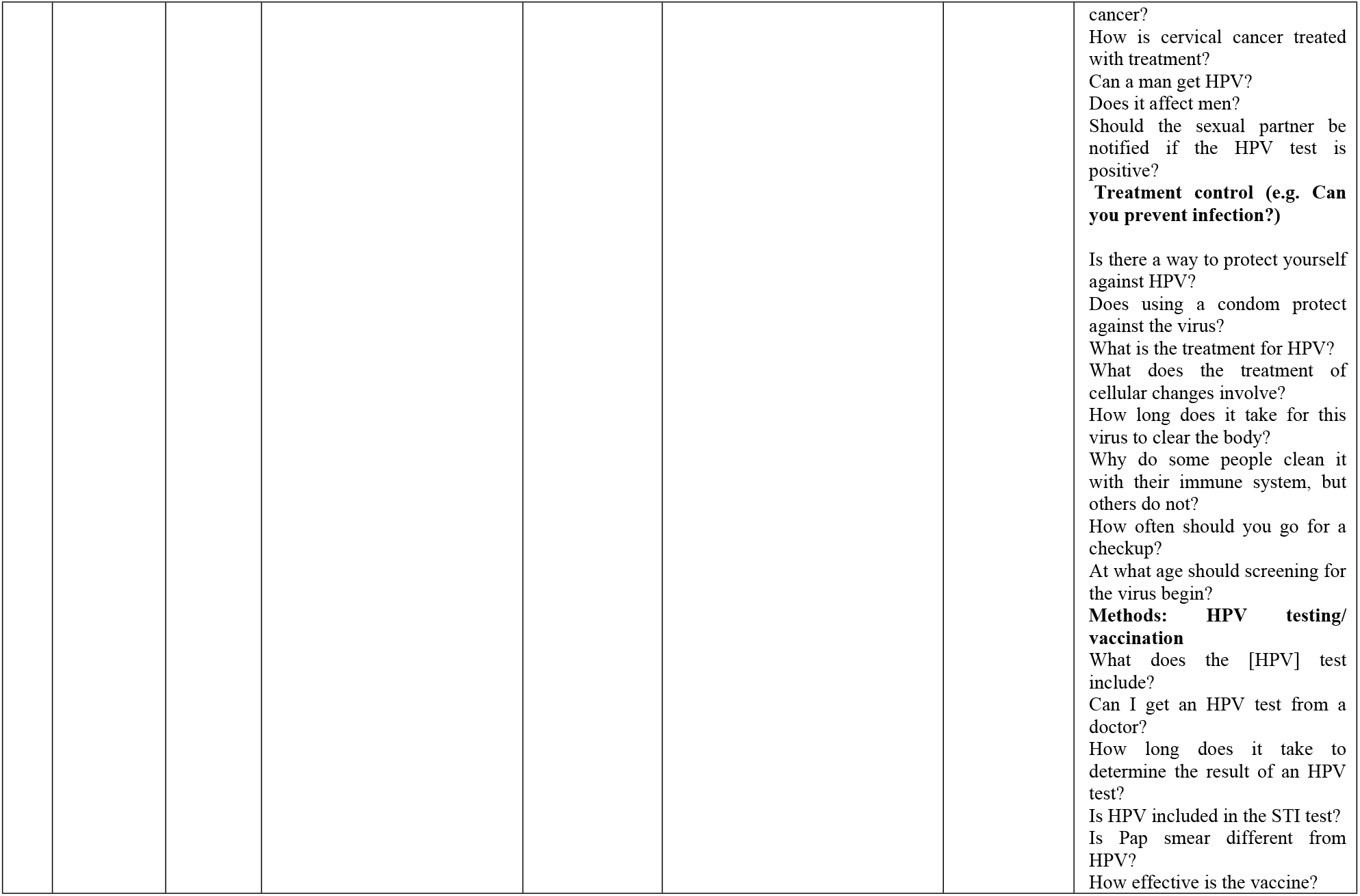
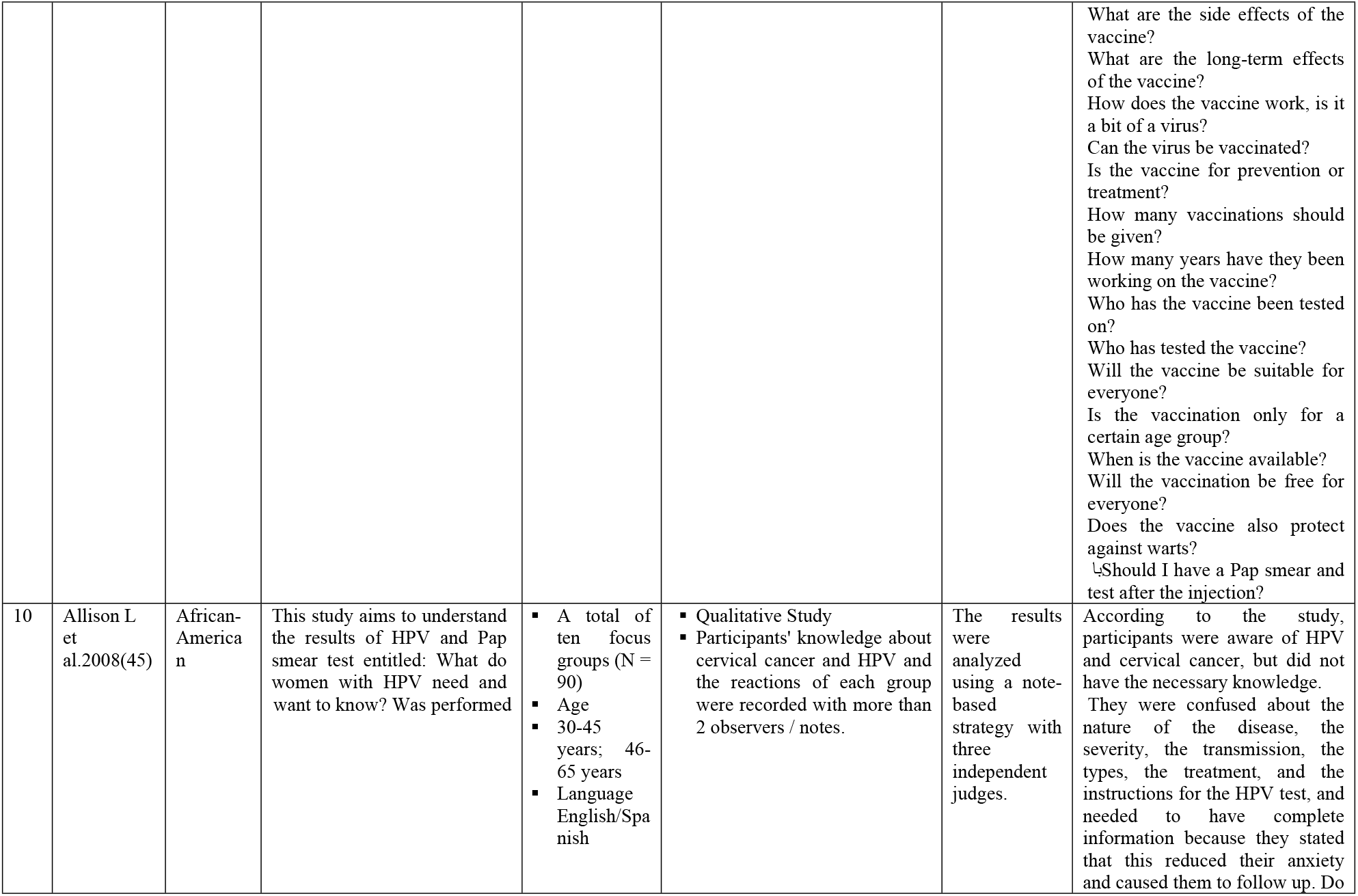
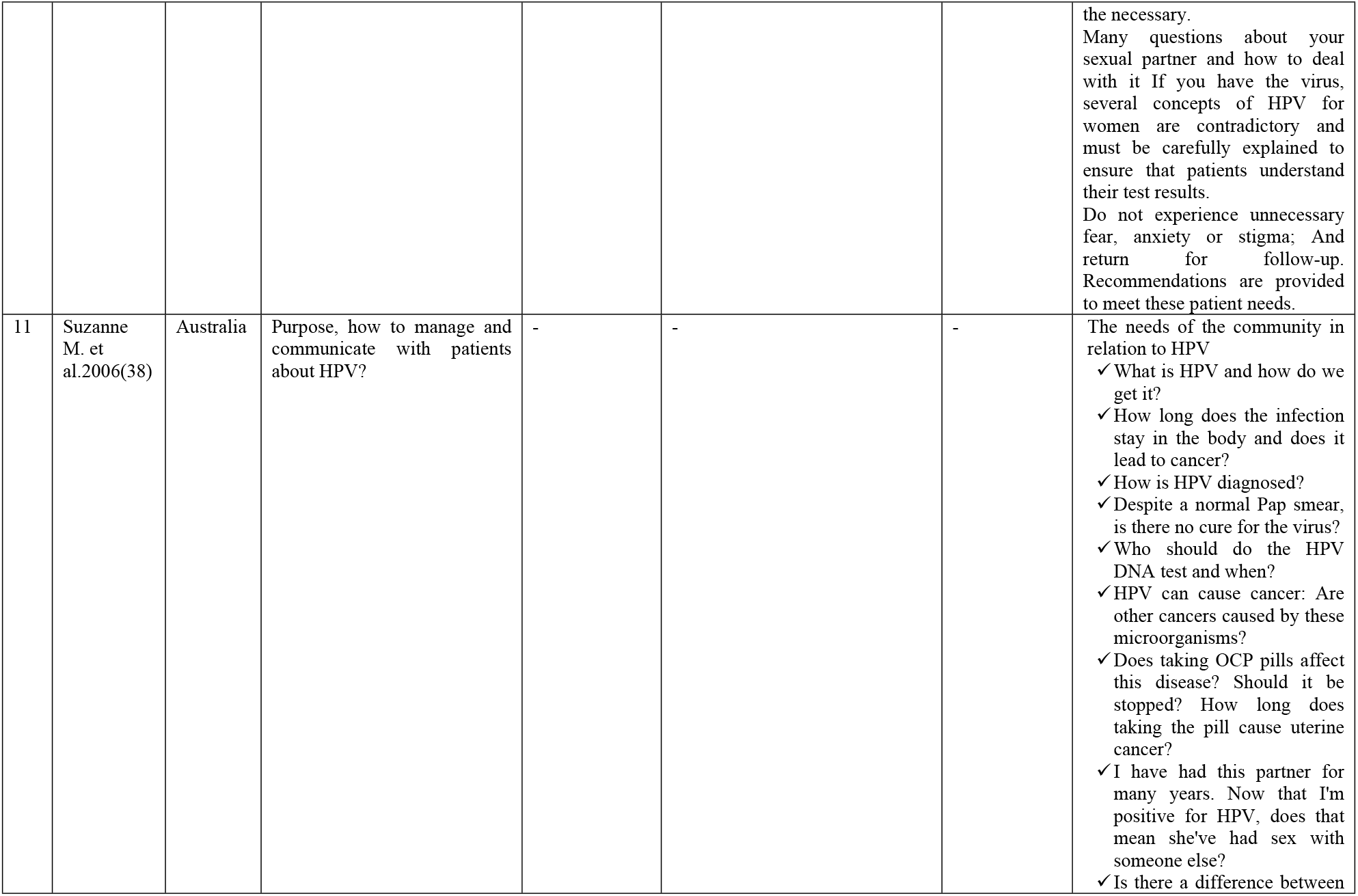
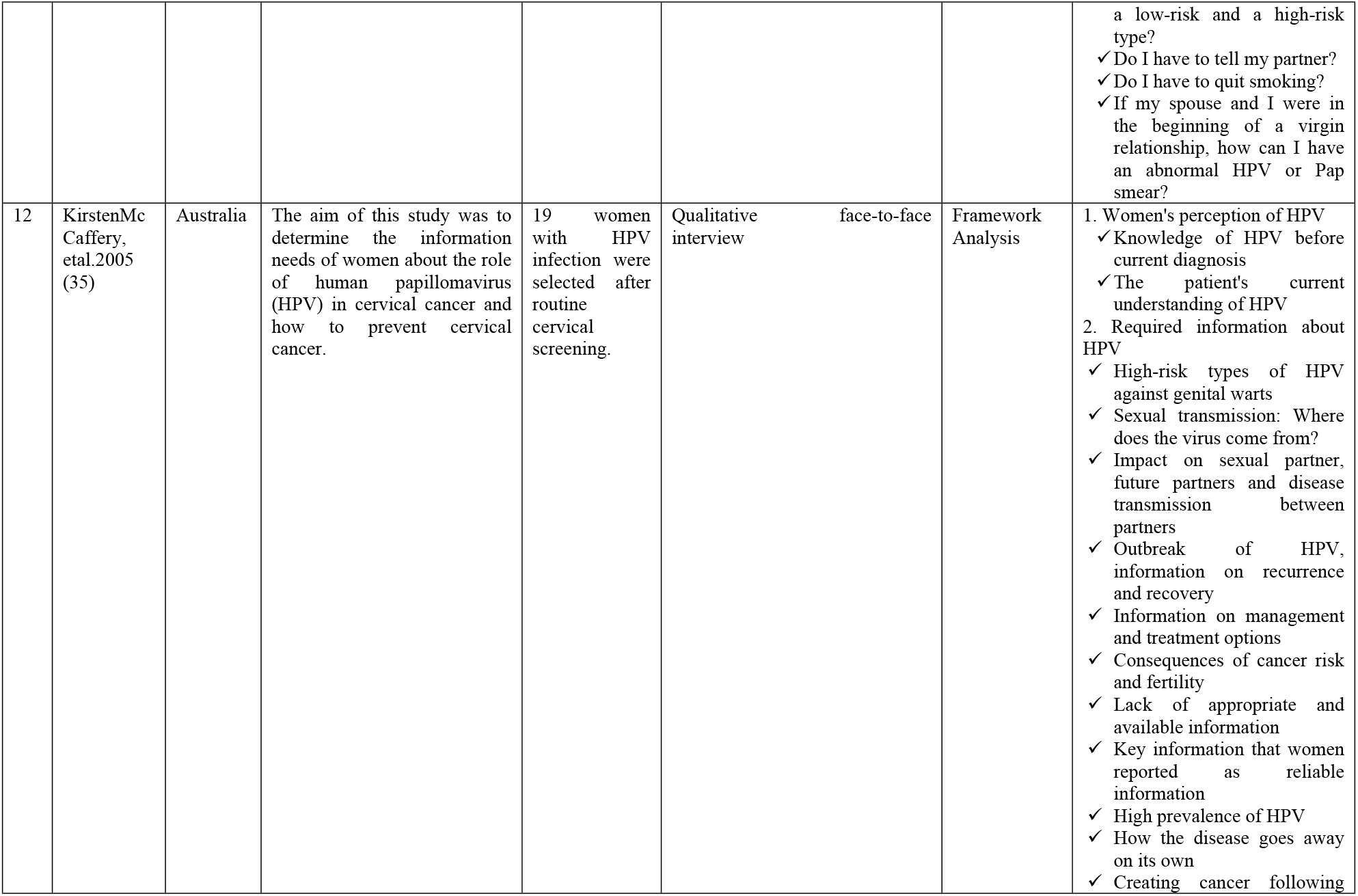
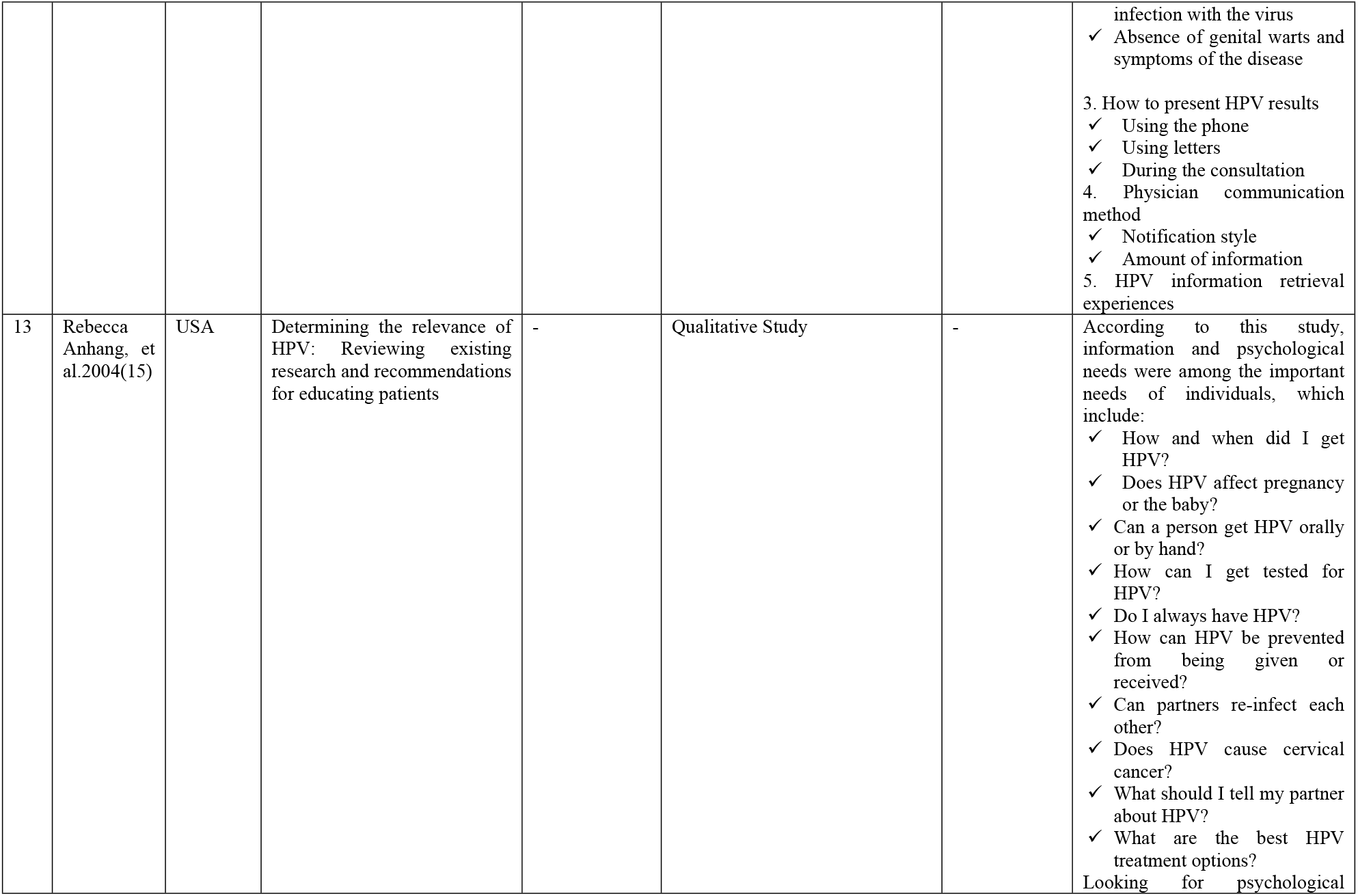
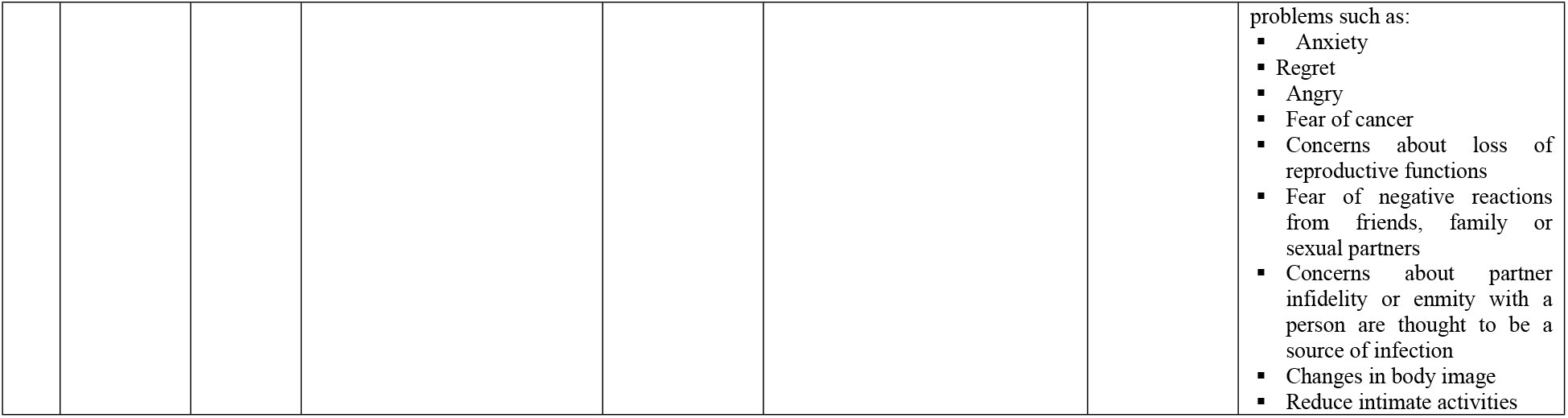
Characteristics of studies in the review

The results of the overall quality evaluation of these articles showed that 8 articles were of good quality, 2 were of average quality and one was of poor quality. There is not enough information about Friedman AL etal and Garland SM articles to evaluate the quality, but since these studies were valuable, they were included in the study according to the opinion of the research team.

The results of the article evaluation are divided into four areas of the checklist, the total score of each article, and article quality in (Tables 2 and 3).

**Table2:**
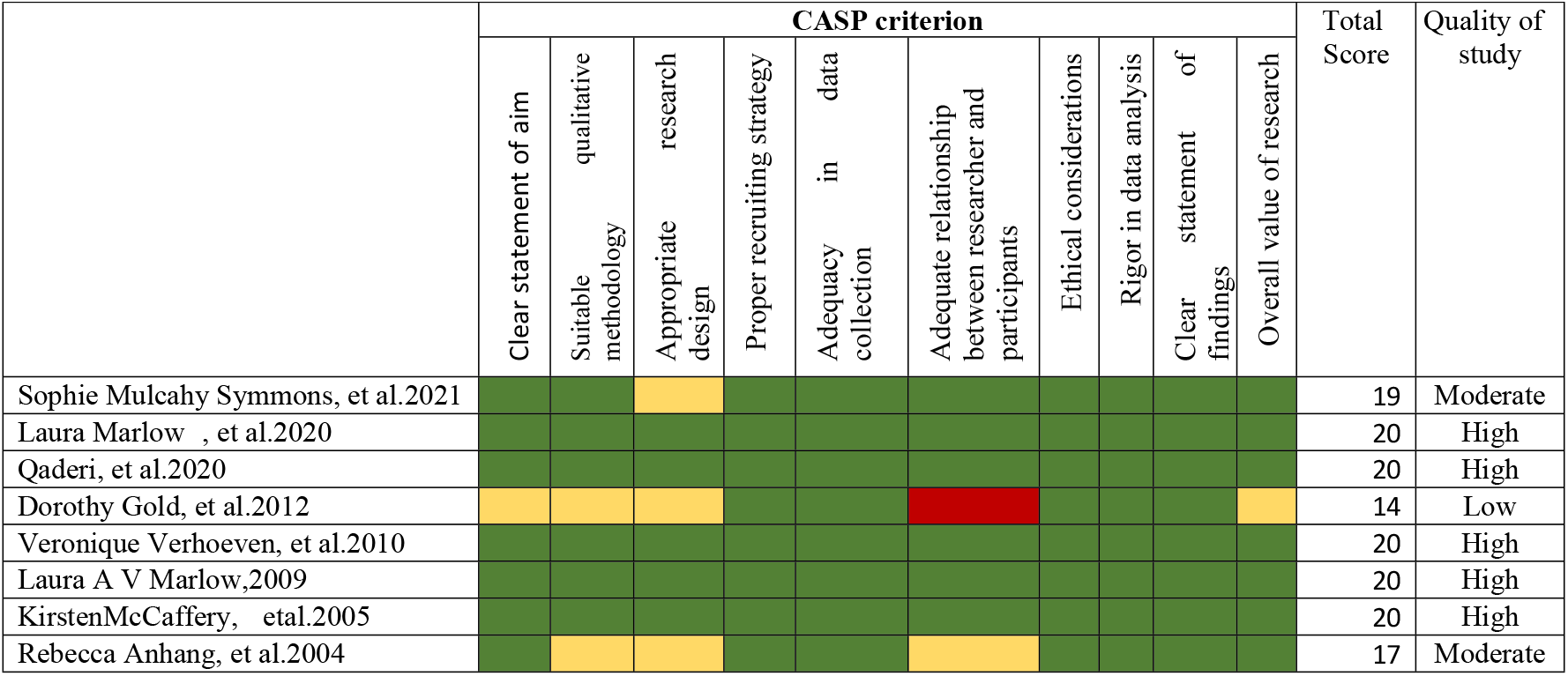
CASP (Critical Appraisal Skills Programme) qualitative checklist scores for total included studies.

**Table3:**
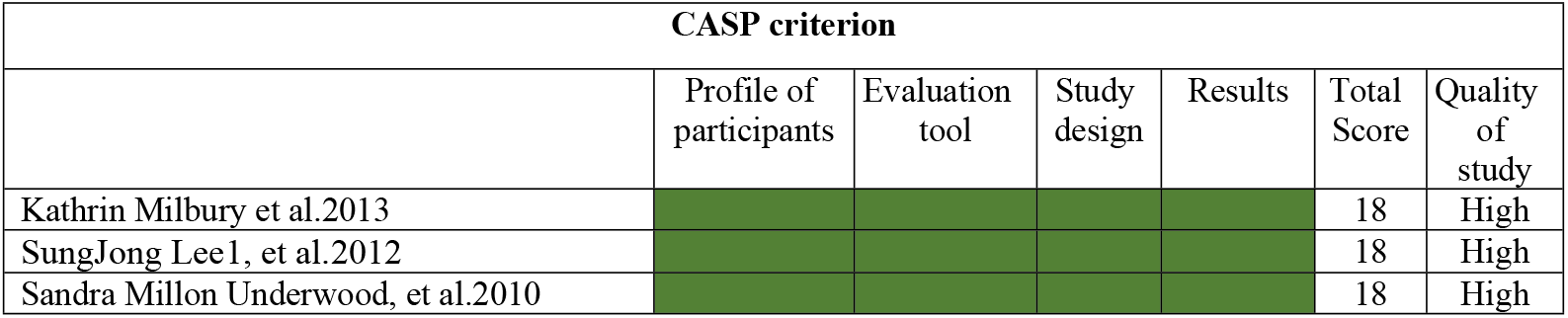
CASP (Critical Appraisal Skills Programme) Crossectional checklist scores for total included studies.

**Figure 1:**
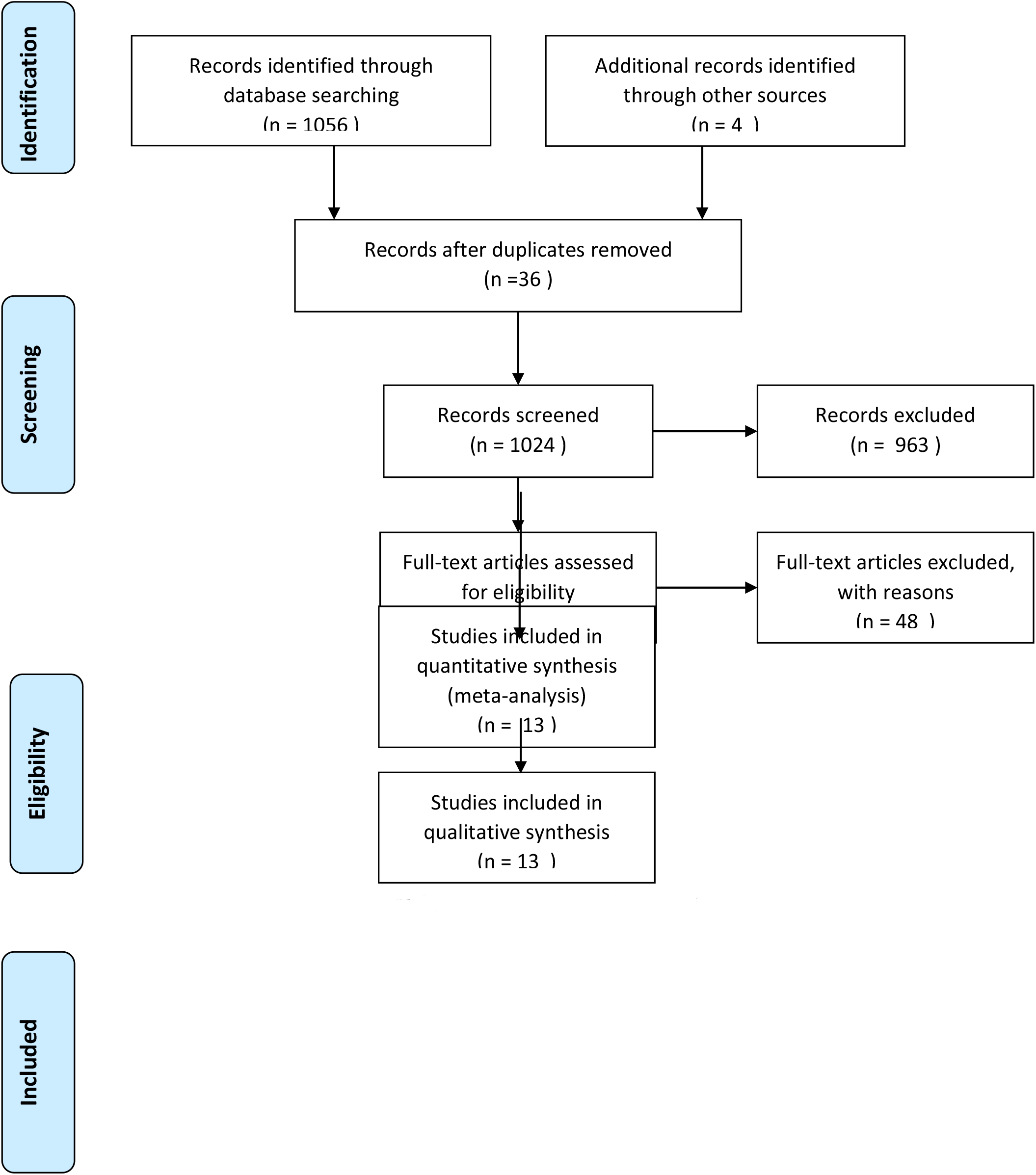
Flow diagram of study selection

Summary of the needs presented by HPV women in qualitative studies including:

People with the virus cognition about HPV and cervical cancer, but did not have the necessary knowledge and awareness about it (26). They were confused about the nature of the disease, its severity, transmission, types, treatment, and instructions for the HPV, and needed to have complete information (27). Therefore, it is necessary to help patients understand the results of their test and their importance. Do not experience unnecessary fear, anxiety, or stigma and seek follow-up and recommendations to meet the patient’s needs.

### Signs, Symptoms, Transmitted, Cause, Consequences of HPV

People with the cause of HPV and how it is transmitted, the signs and symptoms of HPV infection and cervical cancer, how to get screened for cervical cancer, the consequences of the virus and its effects (*for example, does the virus always cause cervical cancer?), Timeline (for example, how long does HPV cause cervical cancer?), Control and treatment (can it be prevented?), Information on how long the infection stays in the body, And does it lead to cancer?* They needed to have complete information because they had stated that it would reduce their anxiety (25, 28-30).

### HPV and sexual partner

Women with HPV needed counseling on how to communicate and deal with a sexual partner. Because some women expressed concern about their husbands’ infidelity and a lack of trust was expressed between the couple (31).

*When I got HPV, How do I tell my partner what to say? What effect does HPV have on sexual life?* Because sometimes these issues lead to uncertainty and stress and in severe cases lead to the breakdown of relationships (28).

The most common question asked by women with HPV was: *which sexual partner (active or former) did the infection come from? Did my partner get it from a previous relationship or did he or she betray me? When HPV is diagnosed in a couple, what are the consequences for the spouse and sexual partner? Will her sexual partner get it too? Should her partner test and follow up? When a couple is infected with HPV and is being treated, will they get the virus again from their partner? Will they constantly infect each other? Will they use a condom? Can I have sex? Can a person get HPV orally or by hand?* (32, 33).

### Seeking needed information by women with HPV

Because many women with the condition felt that they did not have enough information about HPV, many sought more information, including the Internet (social media, blogs, and websites provided by labs and private professionals) (34). Their health care providers and other women living with HPV). They found finding information reliable and up-to-date challenging. Most women prefer the information provided by official websites. Women who were regular Internet users reported finding useful information (31). However, it was often not easy for them to interpret and determine the correctness or incorrectness of the material. Women reported that most cases of finding information about HPV, along with the effect of the virus on genital warts and other sexually transmitted diseases, in which risk factors such as having multiple sexual partners were highlighted, some women were very upset and experienced a sense of shame and embarrassment (35). Or, in many cases, this information was in addition to information related to uterine cancer, which led to fear, anxiety, and stress in them. It seems that the information that women are looking for and the way their doctor informs them can affect their psychological response to HPV infection and is one of the issues that is considered important in follow-up (36).

### HPV and social consequences

Dysfunction or inability to perform daily activities, limitation in relationships with others, limitation of social activities, shock, insecurity, and fear of deformity of the limbs following the development of genital warts and its impact on quality of life were among the challenges (37). Women with HPV experienced it during the illness. Therefore, women with HPV need psychosocial care at different stages of the disease, so that non-fulfillment of psychosocial needs can complicate the course of treatment and recovery (38).

### Reproductive health needs in women with HPV

Women with HPV raised some issues related to fertility, and the need for proper counseling on the impact of the virus on reproductive health was felt (34).

Many women reported seeking information about fertility and pregnancy from various sources. Many women seek information about the adverse effects of HPV on male fertility, the negative impact of HPV on female fertility, the effect of treatments and vaccinations on female fertility and its relationship to female fertility, maternal health threats during pregnancy, adverse pregnancy outcomes, fetal harm, Fear of having a newborn This fear was so great that some people who decided to become pregnant postponed their pregnancy program until the cytology results improved so that they could receive appropriate counseling during this time. Women also noted the association between genital HPV infection and various maternal-fetal variables and pregnancy complications such as miscarriage and preterm delivery Therefore, these women need to know what to do to protect their reproductive health and reduce its complications against infection (30).

Since genital warts can multiply during pregnancy, removing warts during pregnancy was one of the most important issues. Women with genital warts have revealed the need for information about the teratogenicity of some wart removal treatments. They were anxious about any threat it might pose to the fetus (37).

Information on contraception, fear of premature menopause, fear of cervical cancer, fear of hereditary cancer caused by HPV were other issues raised in this area (30).

To prevent unplanned pregnancies, women with HPV needed more information to choose the preferred method of contraception. Because long-term use of birth control pills increases the risk of cervical cancer for women with persistent HPV, they needed guidance in changing their method of contraception. Women sought information on the negative effects of combined oral contraceptive pills (COCs) and levonorgestrel (LNG) pills on their cellular changes (39).

### HPV screening and cervical cancer and vaccinations

Another piece of information that infected women needed to know about was screening. They needed to know how helpful these screenings can be in diagnosing uterine cancer (37). These include: *How is screening done? Is a Pap smear enough to diagnose? Can HPV be diagnosed before cancer develops? In addition to annual checkups, what type of checkups are started at what intervals and at what age? What does the [HPV] test include? Can I get an HPV test from a doctor? How long does it take to determine the result of an HPV test? What is the difference between an HPV test and a Pap smear?(28)*

The women also asked for information about the effect of the vaccine, side effects, long-term effects, mechanism of action of the vaccine, number of doses received, and the age range for vaccination (29).

## Quantitative studies

SungJong Lee, et al. In their quantitative study showed 10 categories for topics and sample questions with different topics. Of the 3,062 people who visited the HPV website, 2,330 asked general questions and 732 asked private questions. The type and frequency of public and private questions showed a statistical difference between the two groups (p<0.001).

From 2004 to 2011 the most common questions, respectively: a. Treatment of HPV and cervical dysplasia (37.8) 1156, b. How HPV transmission (22.3) 684, c. HPV regression (15.7) 481, d. Genital warts (5.7) 174, e. Risk factors for cervical cancer (4.8) 147, f. HPV vaccination 116 (8.3), g. Prevention of HPV 90 (9.2), h. Vaginal discharge (2.8) 86, i. HPV and pregnancy (2.6) 81, j. Laryngeal cancer and HPV 47 (1.5) was (37).

Sandra Millon Underwood et al. showed that sexual behavior and chronic HPV infections are risk factors for cervical cancer. Sexual behavior and chronic HPV infections are risk factors for cervical cancer. When asked to describe their sexual activity, 76.8% (352) of women reported having had sexual activity in the past six months. Among them, 2.62 (219) reported, having a sexual partner, 15.6% (55) two sexual partners and 22.2% (78) three or more partners in the past six months. The most common questions included: cause of HPV, cause of cervical cancer, signs, and symptoms of HPV infection, signs and symptoms of cervical cancer, HPV and cervical cancer screening, HPV vaccine, HPV and cervical cancer risk management (28).

Kathrin Milbury, et,al in their study showed that 66% of patients were aware of their HPV status, but only 35 percent identified HPV as a possible cause of their cancer. Most patients disclosed their HPV status to their partner, 41% talked about transmitting the virus, and only 23% felt aware of the potential risks of transmission and precautions. 39% of their oncologists want to discuss more HPV issues, and 58% sought it from other sources. More than a third said they were interested in more information about HPV.

Women are interested in receiving any information. 18% want more information on how HPV causes cancer. 15% wanted information on HPV vaccination (especially whether vaccinating their children to prevent cancer), 10% wanted information on how to prevent transmission to their partner, and 10% wanted to know if there was any cure for Is there HPV or not? (33).

## Discussion

Based on the review, this systematic review is one of the few review studies that comprehensively addresses the needs of women with HPV. The potential advantage of this study is that the HPV-related questions and needs raised by patients were collected. Recently, many researchers have used written surveys, interviews, and other data collection methods to assess knowledge about HPV among women. They found that women have relatively little knowledge of HPV and suggested using more training programs (40, 41). In addition, many patients become confused after hearing their doctor’s description of HPV. As a result, many patients search the Internet for seeking information about HPV, but there are still many questions for them (42). According to the findings, most people, especially when it comes to frequently asked questions, really want to find out about the whole disease process, from HPV infection to recovery (35). Since most of the studies studied were qualitative studies, this study was able to identify a wide range of needs of affected women. The needs of women with HPV lead to fear, anxiety and worry. Their anxiety is largely due to their poor knowledge of the virus. Therefore, women who are screened for or exposed to the virus also need quality information about HPV and its role in cervical cancer when faced with an abnormal result.

Physicians potentially play an important role in modulating the effects of diagnosis through how HPV is diagnosed (43). Therefore, recognizing these needs can help health professionals understand what questions they are expected to answer.

KirstenMcCaffery and Les Irwig showed in her study that women needed more information about different types of HPV, transmission, effects of the virus on sexual partners, prevalence, delay and regression of HPV, management options, and infection consequences for cancer and fertility risk. Uncertainty about key aspects of HPV, how the physician announced the outcome, and how the outcome was presented (letter, telephone, or consultation) affected the psychological response of women with HPV (35).

Women’s experience of searching the Internet for more information on HPV has been reported as problematic, anxious, and stigmatizing infection in the community, as the information provided is often related to other sexually transmitted infections and multiple sexual partners as a risk factor for Infections are prominent (44). Sung-Jong Lee shows in his study that when people go to a website about HPV, they are looking for “HPV treatment and cervical dysplasia”, “HPV transmission”, “HPV cure”, “Genital warts” And “risk of cervical cancer.” Also, our results showed that “genital warts” and “HPV and pregnancy” may be considered embarrassing topics. Therefore, these findings can be used to provide informed recommendations for clinical or Internet-based communication with patients and the general public (37).

Sandra Millon Underwood In reviewing the questions reported by women with HPV, mentioned five related topics such as: cervical cancer, HPV/cervical cancer prevention, and risk management (28).

Numerous questions asked by women focused on the origin of HPV, the different types of HPV, HPV and risk factors for cervical cancer, the risk factors associated with HPV transmission, and the development of cervical cancer. Similar questions were asked about “early signs”, “warning signs”, symptoms associated with HPV infection and cervical cancer, and if, when and how often women should be tested. However, most of the questions asked by women focused on HPV and cervical cancer prevention and risk management. Among them were specific questions about how HPV was transmitted. Questions about the proper use and effectiveness of male and female inhibitory methods (eg, condoms) to reduce the risk of HPV and cervical cancer; Questions about the safety, availability and cost of the HPV vaccine; and questions about the need for further community education and information (28).

## Conclusion

Surveys showed that the majority of women had unanswered questions about their HPV test results. The information that women thought helped interpret their test results included having a high-risk type of HPV, the risk of short-term and long-term cancer, and cancer survival statistics for the virus. Women also needed information about sexual transmission, how HPV tested positive in a long-term relationship, and the potential consequences for their partners and the risk of re-infection. Younger women had questions about whether HPV could affect fertility.

HPV information should be scientifically accurate and minimize any possible stigma that may be associated with the infection due to its sexually transmitted nature. It was found that seeking information from alternative sources also increased women’s anxiety by providing information about HPV in the context of other highly sexually transmitted diseases and highlighting several sexual partners as a risk factor for infection.

The strength of this study is that it was a comprehensive and systematic review using PRISMA guidelines. In addition, an extensive search strategy was performed without any history or language restrictions. It is possible that due to the range of terms that can be used to describe the needs, some eligible studies may not have been identified in our search, however, we did forward and backward citation searching to reduce this possibility. The data were also extracted by one author and reviewed independently by the second author. While qualitative synthesis allows us to identify a range of different factors that help women with HPV. However, since the majority of studies were qualitative, meta-analysis was not possible. It is suggested that future studies should measure the effect of raising awareness about HPV over time to determine whether raising awareness leads to improved health of infected women. Knowing when and how this response to needs is most effective can help determine the most appropriate interventions.

## Data Availability

..

## Abbreviations

HPV: Human Papillomavirus
COCs: contraceptive pills
LNG: levonorgestrel

## Declarations

### Details of Ethics Approval

This study has been performed in accordance with the Declaration of Helsinki and has been reviewed and approved by the Ethics Committee of Shahroud University of Medical Sciences The: (IR.SHE.REC.1400.154) and after receiving the code from the Prospero system Code: CRD42021293223 Based on the proposed systematic review and meta-analysis checklist (PRISMA) was done.

### Consent of participate

Not applicable

### Consent for publication

Not applicable

### Availability of data and materials

The data that support the findings of this study are available from the corresponding author, [MG], upon reasonable request.

## Acknowledgments

Not applicable

## Authors’ Contributions

MG, ZM,SHZ,HSH designed the studyHSH and ZM planned and undertook the analysis. MG,ZM, and HSH wrote the initial and subsequent drafts of the manuscript. MG,SHZ,ZM and HSH contributed to revising the manuscript. All authors read and approved the final manuscript.

